# COVID-19 Vaccine Effectiveness against the Omicron BA.2 variant in England

**DOI:** 10.1101/2022.03.22.22272691

**Authors:** Freja C. M. Kirsebom, Nick Andrews, Julia Stowe, Samuel Toffa, Ruchira Sachdeva, Eileen Gallagher, Natalie Groves, Anne-Marie O’Connell, Meera Chand, Mary Ramsay, Jamie Lopez Bernal

## Abstract

The BA.1 sub-lineage of the Omicron (B.1.1.529) variant, first detected in the UK in mid-November 2021, rapidly became the dominant strain partly due to reduced vaccine effectiveness. An increase in a second Omicron sub-lineage BA.2 was observed in early January 2022. In this study we use a test-negative case control study design to estimate vaccine effectiveness against symptomatic disease with BA.1 and BA.2 after one or two doses of BNT162b2, ChAdOx1-S or mRNA-1273, and after booster doses of BNT162b2 or mRNA-1273 during a period of co-circulation. Overall, there was no evidence that vaccine effectiveness against symptomatic disease is reduced following infection with the BA.2 sub-lineage as compared to BA.1. Furthermore, similar rates of waning were observed after the second and booster dose for each sub-lineage. These data provide reassuring evidence of the effectiveness of the vaccines currently in use against symptomatic disease caused by BA.2.

## Background

The Omicron (B.1.1.529) variant, first detected in the UK in mid-November 2021, rapidly became the dominant strain partly due to reduced vaccine effectiveness (VE) [1]. An increase in sequenced cases of the Omicron sub-lineage BA.2 was observed in early January 2022 [2]. BA.2 does not contain the spike gene deletion and can be distinguished from BA.1 on whole genome sequencing and on S-gene target status.

BA.2 has a growth advantage over BA.1 [3, 4] and has become the dominant strain in the UK and Denmark. Neutralisation assays using monoclonal antibodies have suggested a small difference between BA.1 and BA.2, although sera from individuals with booster vaccinations neutralise both variants similarly [3].

The UK COVID-19 vaccination program has been in place since December 2020 with primary courses of two doses of either BNT162b2 (Pfizer-BioNTech, Comirnaty^®^), ChAdOx1-S (Vaxzevria, AstraZeneca) or mRNA-1273 (Spikevax, Moderna). Booster vaccination with either BNT162b2 or a half dose (50µg) of mRNA-1273 was introduced in September 2021 to adults over 50 years old and those in risk groups, and in November 2021 expanded to all adults.

In this study we estimate VE against symptomatic disease with BA.1 and BA.2 after one or two doses of BNT162b2, ChAdOx1-S or mRNA-1273, and after booster doses of BNT162b2 or mRNA-1273 during a period of co-circulation.

## Methods

A test-negative case-control study design was used to estimate VE against PCR-confirmed symptomatic COVID-19 following infection with the BA.1 and BA.2, as previously described [5-8]. Logistic regression was used, with the PCR test result as the dependent variable and cases being those testing positive (stratified in separate analyses as either BA.1 or BA.2) and controls being those testing negative. Analysis combined all vaccine manufacturers. Vaccination status was included as an independent variable and effectiveness defined as 1-odds of vaccination in cases/odds of vaccination in controls (full details in Supplementary Appendix).

## Results

Between 17 January 2022 and 17 February 2022 there were a total of 626,148 eligible tests in those aged 18 years and older, with a test date within 10 days of the symptom onset date and which could be linked to the National Immunisation Management system. Of these, 214,171 were BA.1 cases, 31,238 were BA.2 cases and 380,739 were negative tests (controls). A description of eligible tests is included in Supplementary Figure 1 and Supplementary Table 1.

Overall, there was no evidence that VE against symptomatic disease differs between the BA.1 and BA.2 sub-lineages (Figure 1 and Supplementary Table 2). Amongst those who received two doses of any vaccine, VE against symptomatic disease was 63.6% (58.8-67.8%) and 67.1% (54.2-76.3%) for BA.1 and BA.2 respectively within the first two weeks of receiving the second dose. This dropped to 17.4% (15.2-19.4%) and 24.3% (20.3-28.0%) after 25 or more weeks for BA.1 and BA.2, respectively. Amongst those who received any booster dose following immunisation with a primary course of any vaccine, VE increased to 71.3% (69.6-72.9%) and 72.2% (67.0-76.5%) for BA.1 and BA.2 respectively, after a week. Over time, this waned to 45.5% (43.8-47.2%) and 48.4% (45.2-51.4%), respectively, at 15 or more weeks after receiving the booster dose.

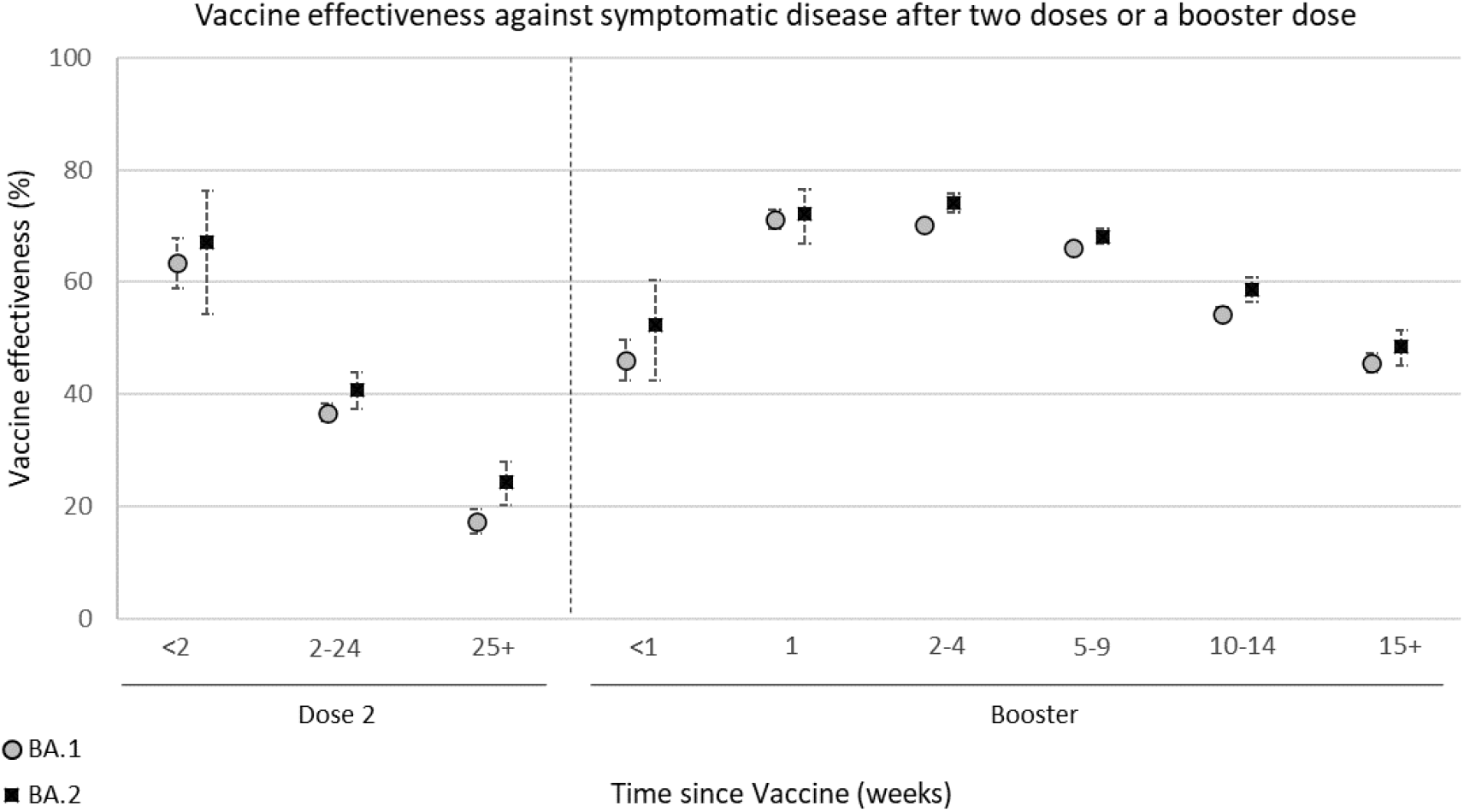

## Discussion

Here we find no reduction in VE against symptomatic disease with BA.2 as compared to BA.1. As previously observed, we find that VE wanes over time [1, 8-10] but there was no difference in the rate of decline. Due to low numbers of BA.2 cases we were not able to stratify by manufacturer in these analyses, however, we found little difference by vaccine after a third dose in previous analyses of the Omicron variant [1].

These findings are consistent with neutralisation assays [3], but there is discrepancy with a Danish household transmission study which found that BA.2 was associated with increased susceptibility to infection for vaccinated individuals [4]. Differences between the UK and Denmark may be explained by different vaccination and infection histories, or by methodological differences, and needs further exploration.

VE against severe disease is higher and retained for longer than effectiveness against mild disease for Omicron BA.1 [9]. Future studies will be needed to estimate VE against severe disease for BA.2, however, based on these data and on experience with other variants, we expect protection against severe disease to be higher than observed here.

## Data Availability

All data produced in the present work are contained in the manuscript and supplementary appendix.

## Authors’ contributions

FCMK and JLB wrote the manuscript. JLB, NA conceptualised the study. FCMK, JS, ST, RS EG, NG, AO’C curated the data. NA conducted the formal analysis. All co-authors reviewed the manuscript.

## Ethics committee approval

PHE Research Ethics and Governance Group Statement: Surveillance of COVID-19 testing and vaccination is undertaken under Regulation 3 of The Health Service (Control of Patient Information) Regulations 2002 to collect confidential patient information (http://www.legislation.gov.uk/uksi/2002/1438/regulation/3/made) under Sections 3(i) (a) to (c), 3(i)(d) (i) and (ii) and 3(3). The study protocol was subject to an internal review by the PHE Research Ethics and Governance Group and was found to be fully compliant with all regulatory requirements. As no regulatory issues were identified, and ethical review is not a requirement for this type of work, it was decided that a full ethical review would not be necessary.

All necessary patient/participant consent has been obtained and the appropriate institutional forms have been archived.

