## Supplementary Appendix for "COVID-19 Vaccine Effectiveness against the Omicron BA.2 variant in England"

### Supplementary Methods

#### Study Design

A test negative case control design was used to estimate vaccine effectiveness against symptomatic COVID-19 with the Omicron variant compared to the Delta variant in individuals aged 18 years and older. The odds of vaccination in symptomatic PCR positive cases was compared to the odds of vaccination in symptomatic individuals who tested negative for SARS-CoV-2 in England.

#### Data Sources

##### COVID-19 Testing Data

SARS-CoV-2 Testing PCR testing for SARS CoV-2 in England is undertaken by hospital and public health laboratories (Pillar 1), as well as by community testing (Pillar 2). Pillar 2 testing is available to anyone with symptoms consistent with COVID-19 (high temperature, new continuous cough, or loss or change in sense of smell or taste), anyone who is a contact of a confirmed case, care home staff and residents, and to those who have self-tested as positive using a lateral flow test (LFT). Data on all positive PCR and LFTs, and on negative Pillar 2 PCR tests from symptomatic individuals with a test date after 25 November 2020 were extracted up to 17 February 2022. Individuals who reported symptoms and were tested in Pillar 2 between 17 January 2022 and 17 February 2022 were included in the analysis (Supplementary Figure 1). Any negative tests taken within 7 days of a previous negative test, and any negative tests where symptom onset date was within the 10 days or a previous symptoms onset date for a negative test were dropped as these likely represent the same episode. Negative tests taken within 21 days of a subsequent positive test were also excluded as chances are high that these are false negatives. Positive and negative tests within 90 days of a previous positive test were also excluded; however, where participants had later positive tests within 14 days of a positive then preference was given to PCR tests and symptomatic tests. For individuals who had more than one negative test, one was selected at random in the study period. Data were restricted to persons who had reported symptoms and gave a symptom onset date within the 10 days before testing to account for reduced PCR sensitivity beyond this period in an infection event.

##### Vaccination Data

The National Immunization Management System (NIMS) contains demographic information on the whole population of England who are registered with a general practice physician in England and is used to record all COVID-19 vaccinations. NIMS was accessed for dates of vaccination and manufacturer, sex, date of birth, ethnicity, and residential address. Addresses were used to determine index of multiple deprivation quintile and were also linked to Care Quality Commission registered care homes using the unique property reference number. Data on geography (NHS region), risk group status, clinically extremely vulnerable status, and health/social care worker were also extracted from the NIMS. Clinical risk groups included a range of chronic conditions as described in the Green Book, whereas the clinically extremely vulnerable group included persons who were considered to be at the highest risk for severe COVID-19, including those with immunosuppressed conditions and those with severe respiratory disease. Booster doses were identified as a third dose given at least 84 days after a second dose and administered after 13 September 2021. Individuals with four or more doses of vaccine, heterologous primary schedule or fewer than 19 days between their first and second dose were excluded.

Testing data were linked to NIMS on 8 February 2021 using combinations of the unique individual National Health Service (NHS) number, date of birth, surname, first name, and postcode using deterministic linkage – 99.6% of eligible tests could be linked to the NIMS.

### Identification of Delta and Omicron Variants and assignment to cases

Sequencing of PCR positive samples is undertaken through a network of laboratories, including the Wellcome Sanger Institute. Whole-genome sequences are assigned to UKHSA definitions of variants based on mutations. S-gene target status on PCR-testing is an alternative approach for identifying each sub-lineage because BA.1 has been associated with S-gene target failure on PCR testing with the Taqpath assay while BA.2 has been associated with a positive S-gene target. Cases were defined as BA.1 or BA.2 based on whole genome sequencing or S-gene target status, with sequencing taking priority. Where subsequent positive tests within 14 days included sequencing or S-gene target failure information, this information was used to classify the variant.

### Statistical Analysis

Logistic regression was used, with the PCR test result as the dependent variable and cases being those testing positive (stratified in separate analyses as either BA.1 or BA.2) and controls being those testing negative. Vaccination status was included as an independent variable and effectiveness defined as 1- odds of vaccination in cases/odds of vaccination in controls.

Vaccine effectiveness was adjusted in logistic regression models for age (ages 18-19, then 20 through to 89 in five-year bands, then everyone age 90 years or older), sex, index of multiple deprivation (quintile), ethnic group, history of travel, geographic region (NHS region), period (week of test), health and social care worker status, clinical risk group status, clinically extremely vulnerable, and previously testing positive. These factors were all considered potential confounders so were included in all models.

Analysis combined all vaccine manufacturers (ChAdOx1, BNT162b2 or mRNA-1273 for one dose or two doses, and BNT162b2 or mRNA-1273 (half-dose) for booster doses). Heterologous primary schedules and ChAdOx1 primary course followed by ChAdOx1 booster dose recipients were excluded.

Vaccine effectiveness was assessed in intervals of <4 weeks and 4+ weeks post the first dose; <2 weeks, 2-24 and 25+ weeks post the second dose; and < 1 week, 1, 2-4, 5-9, 10-14 and 15+ weeks post a booster dose.

### Supplementary Tables and Figures

#### Supplementary Figure 1.

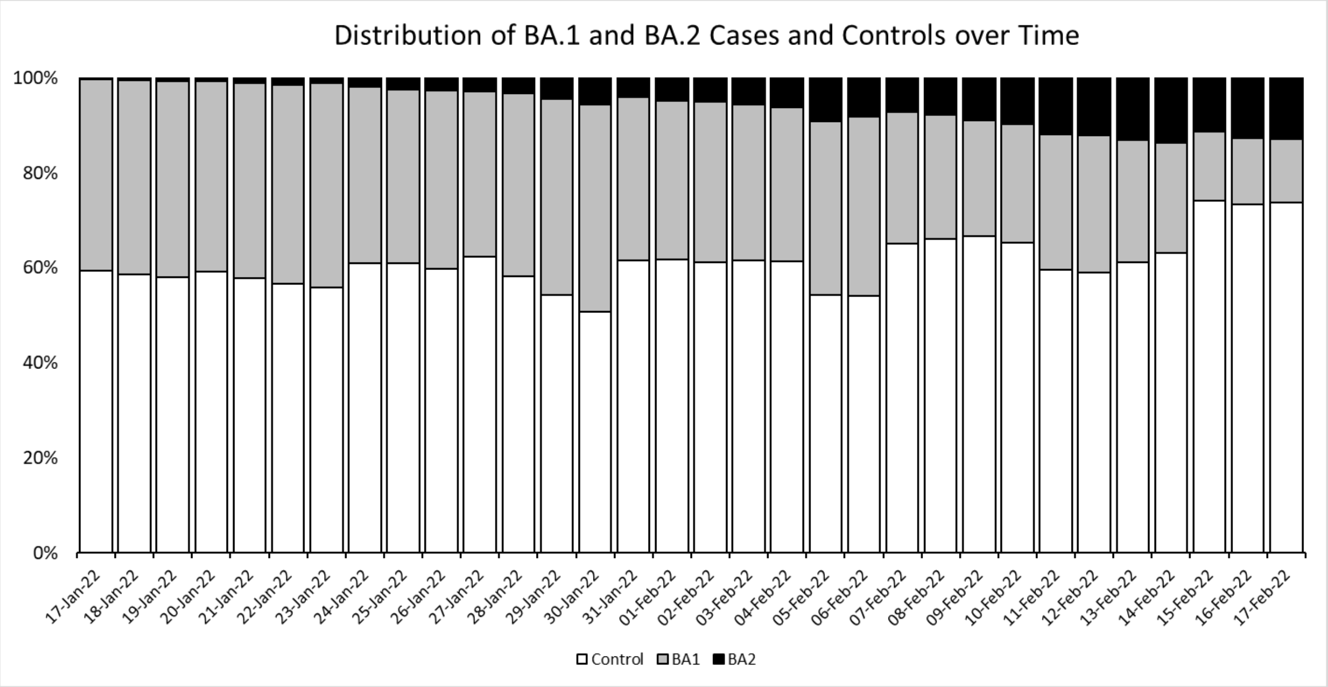

### Supplementary Table 1

| | | | Overall | | Negative | | BA.1 | | BA.2 | |
|---|---|---|---|---|---|---|---|---|---|---|
| | | | n | % | n | % | n | % | n | % |
| | Test Result | Interval (weeks) | **626,148** | **100.0%** | **380,739** | **60.8%** | **214,171** | **34.2%** | **31,238** | **5.0%** |
| **Vaccination Status and intervals after vaccine** | Unvaccinated | | 56,656 | **9.0%** | 23,839 | **6.3%** | 28,589 | **13.3%** | 4,228 | **13.5%** |
| | Dose 1* | <4 | 3,048 | **0.5%** | 1,754 | **0.5%** | 1,147 | **0.5%** | 147 | **0.5%** |
| | | 4+ | 16,503 | **2.6%** | 9,155 | **2.4%** | 6,422 | **3.0%** | 926 | **3.0%** |
| | Dose 2* | <2 | 1,331 | **0.2%** | 900 | **0.2%** | 392 | **0.2%** | 39 | **0.1%** |
| | | 2-24 | 55,941 | **8.9%** | 30,431 | **8.0%** | 22,649 | **10.6%** | 2,861 | **9.2%** |
| | | 25+ | 58,630 | **9.4%** | 27,191 | **7.1%** | 27,676 | **12.9%** | 3,763 | **12.0%** |
| | Booster** | <1 | 4,189 | **0.7%** | 2,375 | **0.6%** | 1,681 | **0.8%** | 133 | **0.4%** |
| | | 1 | 7,074 | **1.1%** | 5,009 | **1.3%** | 1,917 | **0.9%** | 148 | **0.5%** |
| | | 2-4 | 71,415 | **11.4%** | 50,272 | **13.2%** | 19,690 | **9.2%** | 1,453 | **4.7%** |
| | | 5-9 | 211,960 | **33.9%** | 144,591 | **38.0%** | 57,939 | **27.1%** | 9,430 | **30.2%** |
| | | 10-14 | 93,988 | **15.0%** | 58,191 | **15.3%** | 30,881 | **14.4%** | 4,916 | **15.7%** |
| | | 15+ | 45,413 | **7.3%** | 27,031 | **7.1%** | 15,188 | **7.1%** | 3,194 | **10.2%** |
| **Age** | 18-19 | | 17,575 | **2.8%** | 11,215 | **2.9%** | 5,583 | **2.6%** | 777 | **2.5%** |
| | 20-24 | | 54,760 | **8.7%** | 33,884 | **8.9%** | 17,944 | **8.4%** | 2,932 | **9.4%** |
| | 25-29 | | 72,059 | **11.5%** | 43,679 | **11.5%** | 24,504 | **11.4%** | 3,876 | **12.4%** |
| | 30-34 | | 89,635 | **14.3%** | 54,422 | **14.3%** | 30,734 | **14.4%** | 4,479 | **14.3%** |
| | 35-39 | | 91,105 | **14.6%** | 54,598 | **14.3%** | 32,297 | **15.1%** | 4,210 | **13.5%** |
| | 40-44 | | 79,118 | **12.6%** | 46,998 | **12.3%** | 28,137 | **13.1%** | 3,983 | **12.8%** |
| | 45-49 | | 62,566 | **10.0%** | 37,779 | **9.9%** | 21,659 | **10.1%** | 3,128 | **10.0%** |
| | 50-54 | | 52,447 | **8.4%** | 32,094 | **8.4%** | 17,720 | **8.3%** | 2,633 | **8.4%** |
| | 55-59 | | 40,650 | **6.5%** | 24,907 | **6.5%** | 13,777 | **6.4%** | 1,966 | **6.3%** |
| | 60-64 | | 27,343 | **4.4%** | 16,879 | **4.4%** | 9,125 | **4.3%** | 1,339 | **4.3%** |
| | 65-69 | | 16,352 | **2.6%** | 10,440 | **2.7%** | 5,090 | **2.4%** | 822 | **2.6%** |
| | 70-74 | | 11,039 | **1.8%** | 6,893 | **1.8%** | 3,632 | **1.7%** | 514 | **1.6%** |
| | 75-79 | | 5,899 | **0.9%** | 3,586 | **0.9%** | 1,989 | **0.9%** | 324 | **1.0%** |
| | 80-84 | | 2,955 | **0.5%** | 1,780 | **0.5%** | 1,031 | **0.5%** | 144 | **0.5%** |
| | 85-89 | | 1,661 | **0.3%** | 1,033 | **0.3%** | 554 | **0.3%** | 74 | **0.2%** |
| | >=90 | | 984 | **0.2%** | 552 | **0.1%** | 395 | **0.2%** | 37 | **0.1%** |
| **Gender** | Female | | 391,984 | **62.6%** | 243,987 | **64.1%** | 129,951 | **60.7%** | 18,046 | **57.8%** |
| | Male | | 233,066 | **37.2%** | 136,056 | **35.7%** | 83,866 | **39.2%** | 13,144 | **42.1%** |
| | Missing | | 1,098 | **0.2%** | 696 | **0.2%** | 354 | **0.2%** | 48 | **0.2%** |
| **Ethnicity** | African | | 8,588 | **1.4%** | 4,850 | **1.3%** | 3,204 | **1.5%** | 534 | **1.7%** |
| | Another Asian background | | 10,003 | **1.6%** | 5,571 | **1.5%** | 3,574 | **1.7%** | 858 | **2.7%** |
| | Another Black background | | 777 | **0.1%** | 430 | **0.1%** | 283 | **0.1%** | 64 | **0.2%** |
| | Another ethnic background | | 5,121 | **0.8%** | 2,976 | **0.8%** | 1,780 | **0.8%** | 365 | **1.2%** |
| | Arab | | 2,930 | **0.5%** | 1,689 | **0.4%** | 1,085 | **0.5%** | 156 | **0.5%** |
| | Bangladeshi | | 5,073 | **0.8%** | 2,726 | **0.7%** | 1,911 | **0.9%** | 436 | **1.4%** |
| | Caribbean | | 4,283 | **0.7%** | 2,162 | **0.6%** | 1,783 | **0.8%** | 338 | **1.1%** |
| | Chinese | | 5,594 | **0.9%** | 3,446 | **0.9%** | 1,763 | **0.8%** | 385 | **1.2%** |
| | Indian | | 25,234 | **4.0%** | 15,457 | **4.1%** | 7,942 | **3.7%** | 1,835 | **5.9%** |

| | | | | | | | | | |
|---|---|---|---|---|---|---|---|---|---|
| | Mixed or multiple ethnicities | 11,849 | **1.9%** | 6,996 | **1.8%** | 4,144 | **1.9%** | 709 | **2.3%** |
| | Pakistani | 15,750 | **2.5%** | 8,239 | **2.2%** | 6,901 | **3.2%** | 610 | **2.0%** |
| | Prefer not to say | 25,081 | **4.0%** | 14,651 | **3.8%** | 9,041 | **4.2%** | 1,389 | **4.4%** |
| | White | 505,864 | **80.8%** | 311,546 | **81.8%** | 170,759 | **79.7%** | 23,559 | **75.4%** |
| | Missing | 1 | **0.0%** | 0 | **0.0%** | 1 | **0.0%** | 0 | **0.0%** |
| **NHS Region** | East of England | 86,213 | **13.8%** | 48,723 | **12.8%** | 31,696 | **14.8%** | 5,794 | **18.5%** |
| | London | 91,746 | **14.7%** | 51,270 | **13.5%** | 31,490 | **14.7%** | 8,986 | **28.8%** |
| | Midlands | 113,031 | **18.1%** | 69,172 | **18.2%** | 39,406 | **18.4%** | 4,453 | **14.3%** |
| | North East | 98,992 | **15.8%** | 55,615 | **14.6%** | 41,025 | **19.2%** | 2,352 | **7.5%** |
| | North West | 80,926 | **12.9%** | 42,000 | **11.0%** | 36,217 | **16.9%** | 2,709 | **8.7%** |
| | South East | 95,901 | **15.3%** | 67,967 | **17.9%** | 22,939 | **10.7%** | 4,995 | **16.0%** |
| | South West | 59,336 | **9.5%** | 45,989 | **12.1%** | 11,398 | **5.3%** | 1,949 | **6.2%** |
| | Missing | 3 | **0.0%** | 3 | **0.0%** | 0 | **0.0%** | 0 | **0.0%** |
| **IMD Quintiles** | 1 | 116,174 | **18.6%** | 61,786 | **16.2%** | 49,562 | **23.1%** | 4,826 | **15.4%** |
| | 2 | 124,346 | **19.9%** | 73,002 | **19.2%** | 44,635 | **20.8%** | 6,709 | **21.5%** |
| | 3 | 127,673 | **20.4%** | 78,564 | **20.6%** | 42,226 | **19.7%** | 6,883 | **22.0%** |
| | 4 | 128,056 | **20.5%** | 81,392 | **21.4%** | 40,121 | **18.7%** | 6,543 | **20.9%** |
| | 5 | 127,664 | **20.4%** | 84,661 | **22.2%** | 36,869 | **17.2%** | 6,134 | **19.6%** |
| | Missing | 2,235 | **0.4%** | 1,334 | **0.4%** | 758 | **0.4%** | 143 | **0.5%** |
| **Vaccine priority groups** | HSCW | 50,363 | **8.0%** | 31,964 | **8.4%** | 16,057 | **7.5%** | 2,342 | **7.5%** |
| | At risk*** | 125,427 | **20.0%** | 76,330 | **20.0%** | 43,411 | **20.3%** | 5,686 | **18.2%** |
| | CEV | 40,342 | **6.4%** | 22,638 | **5.9%** | 15,579 | **7.3%** | 2,125 | **6.8%** |
| **Previously positive** | No | 522,920 | **83.5%** | 306,122 | **80.4%** | 189,168 | **88.3%** | 27,630 | **88.4%** |
| | Yes | 103,228 | **16.5%** | 74,617 | **19.6%** | 25,003 | **11.7%** | 3,608 | **11.6%** |

*Dose 1 and 2 is recipients of ChAdOx1-S, BNT162b2 or mRNA-1273.
**Booster is recipients of BNT162b2 or mRNA-1273 following any primary immunisation course.
***At risk is only those under 65

### Supplementary Table 2

| | | | BA.1 | | BA.2 | |
|---|---|---|---|---|---|---|
| Doses | Interval (weeks) | Controls | Cases | VE (95% CI) | Cases | VE (95% CI) |
| Unvaccinated | | 23,839 | 28589 | baseline | 4228 | baseline |
| Dose 1* | <4 | 1,754 | 1147 | 44.7 (40.1-48.9) | 147 | 44.8 (34.0-53.9) |
| | 4+ | 9,155 | 6422 | 40.7 (38.4-42.9) | 926 | 43.0 (38.3-47.4) |
| Dose 2* | <2 | 900 | 392 | 63.6 (58.8-67.8) | 39 | 67.1 (54.2-76.3) |
| | 2-24 | 30,431 | 22649 | 36.8 (35.1-38.4) | 2861 | 40.7 (37.5-43.8) |
| | 25+ | 27,191 | 27676 | 17.4 (15.2-19.4) | 3763 | 24.3 (20.3-28.0) |
| Booster** | <1 | 2,375 | 1681 | 46.1 (42.4-49.7) | 133 | 52.3 (42.6-60.3) |
| | 1 | 5,009 | 1917 | 71.3 (69.6-72.9) | 148 | 72.2 (67.0-76.5) |
| | 2-4 | 50,272 | 19690 | 70.2 (69.5-71.0) | 1453 | 74.2 (72.4-75.8) |
| | 5-9 | 144591 | 57939 | 66.2 (65.5-66.9) | 9430 | 68.1 (66.7-69.5) |
| | 10-14 | 58191 | 30881 | 54.3 (53.1-55.5) | 4916 | 58.6 (56.4-60.7) |
| | 15+ | 27031 | 15188 | 45.5 (43.8-47.2) | 3194 | 48.4 (45.2-51.4) |

*Dose 1 and 2 is recipients of ChAdOx1-S, BNT162b2 or mRNA-1273.
**Booster is recipients of BNT162b2 or mRNA-1273 following any primary immunisation course.
